# First detection of SARS-CoV-2 genetic material in the vicinity of COVID-19 isolation centre through wastewater surveillance in Bangladesh

**DOI:** 10.1101/2020.09.14.20194696

**Authors:** Firoz Ahmed, Md. Aminul Islam, Manish Kumar, Maqsud Hossain, Prosun Bhattacharya, Md. Tahmidul Islam, Foysal Hossen, Md. Shahadat Hossain, Md. Sydul Islam, Md. Main Uddin, Md. Nur Islam, Newaz Mohammed Bahadur, Md. Didar-ul-Alam, Hasan Mahmud Reza, Md. Jakariya

**Author notes:** Corresponding Author: Dr. Firoz Ahmed Professor and Chairman & Focal Point COVID-19 Diagnostic Lab, Department of Microbiology, Noakhali Science and Technology University, Noakhali-3814, Bangladesh.

## Abstract

In the course of a COVID-19 pandemic, 0.33 million people got infected in Bangladesh, we made the first and successful attempt to detect SARS-CoV-2 viruses’ genetic material in the vicinity wastewaters of an isolation centre i.e. Shaheed Bhulu Stadium, situated at Noakhali. The idea was to understand the genetic loading variation, both temporal and distance-wise in the nearby wastewater drains when the number of infected COVID-19 patients is not varying much. Owing to the fact that isolation center, in general, always contained a constant number of 200 COVID-19 patients, the prime objective of the study was to check if several drains carrying RNA of coronavirus are actually getting diluted or accumulated along with the sewage network. Our finding suggested that while the temporal variation of the genetic load decreased in small drains over the span of 50 days, the main sewer exhibited accumulation of SARS-CoV-2 RNA. Other interesting finding displays that probably distance of sampling location in meters is not likely to have a significant impact on gene detection concentration, although the quantity of the RNA extracted in the downstream of the drain was higher. These findings are of immense value from the perspective of wastewater surveillance of COVID-19, as they largely imply that we do not need to monitor every wastewater system, and probably major drains monitoring may illustrate the city health. Perhaps, we are reporting the accumulation of SARS-CoV-2 genetic material along with the sewer network i.e. from primary to tertiary drains. The study sought further data collection in this line to simulate conditions prevailed in the most of south Asian country and to shed further light on the temporal variation and decay/accumulation processes of the genetic load of the SARS-COV-2.

## 1. Introduction

Severe acute respiratory syndrome coronavirus 2 (SARS-CoV-2) is the strain of coronavirus that causes coronavirus disease 2019 (COVID-19), that are now being frequently reported in specimens collected from the wastewater treatment plants around the world owing to the shredding of the symptomatic/asymptomatic COVID-19 infection (Ahmed et al., 2020; Haramoto et al., 2020; Kumar et al., 2020; La Rosa et al., 2020; Medema et al., 2020; Nemudryi et al., 2020; Or et al., 2020; Randazzo et al., 2020; Rimoldi et al., 2020; Wu et al., 2020b; Wurtzer et al., 2020). In Milano, Italy, ORF1ab assay and N protein assay was used to detect SARS-CoV-2 RNA from untreated wastewater and river water samples (Rimoldi et al., 2020). However, the wastewater surveillance of COVID-19 (WWSoC-19) has been mostly focused on wastewater treatment plants around the globe, and there is slow progress on ambient waters and especially in the sewer system. This is critical because several developing countries like India, Bangladesh, Pakistan, and others do not have proper wastewater systems even in their class-I cities. As a result, the policymakers in these countries are in confusion pertaining to the national scale implementation of WWSoC-19.

Further, while the infectivity issues of SARS-CoV-2 RNA present in the wastewater as well as ambient environmental waters are not yet neglected or proved in the scholarly world, the public around the world are very skeptical about the wastewaters coming from the isolation centers mainly that are not equipped with wastewater treatment systems. There have been some decay reports of genetic loading of SARS-CoV-2 (Ahmed et al., 2020b, Kumar et al., 2020b) in the wastewater system, but sewer systems are not yet investigated. In addition, there is a lack of explicit understanding of either decay or accumulation of Covid-19 genetic material along with the sewer systems with distance and the networking from small drain to larger drains followed by the canal and main sewer system.

Furthermore, it is a general observation that most of the WWSoC-19 studies reported worldwide have either correlated their C_t_-value or detected gene copies with the total infected person in the corresponding monitoring city or country. Uncertainty is very much evident on the average amount of gene copies shredded by an infected person, and its relationship with the number of genes detected during WWSoC-19. While we already know about the variations that exist in the length of viral shedding in wastewater from the various studies (Wu et al., 2020b; Xu et al., 2020), the magnitude of the shedding ranges between 10^2^ and 10^8^ copies of RNA per gram of human waste (Lescure et al., 2020; Pan et al., 2020; Wölfel et al., 2020). The general trend has been to see the fluctuation in the Ct value and then estimate the corresponding increase or decrease of the COVID-19 patient in a given vicinity of treatment plants. However, there has been a complete lack of WWSoC-19 study where infected individuals’ information is known and then a variation in genetic loading has been studied.

Accordingly, we conducted a preliminary detection of SARS-CoV-2 RNA in wastewater samples from the sewage network in the isolation centre at Noakhali, Bangladesh. The aim of the study was to: i) understand the genetic load in the vicinity of the isolation centre with an almost constant number of COVID-19 patients. ii) analyse the distance impact on genetic loading and tracing the decay/accumulation of the same along with the sewage network, iii) and temporal variation in genetic material shedding from the infected patient.

## 2. Material and Methods

### 2.1 Sampling

Wastewater samples were collected from sewage waste tank, passage drain, and toilets near Shaheed Bhulu Stadium at Noakhali, Bangladesh (**Figure 1**). The sampling location for this study was selected based on the fact that Shaheed Bhulu Stadium is the largest detention Centre for COVID-19 patients at Noakhali district, Bangladesh. This facility has been established to accommodate more than two hundred COVID-19 positive patients for isolation purposes but kept around 200 patients all the time during the monitoring period. This preliminary study has been carried out with samples collected from the three different drains that are coming out of the stadium and can be considered as primary drains, which connects to a canal (secondary drainage system) and eventually meets the main sewer system (tertiary drains).

**Figure 1.**
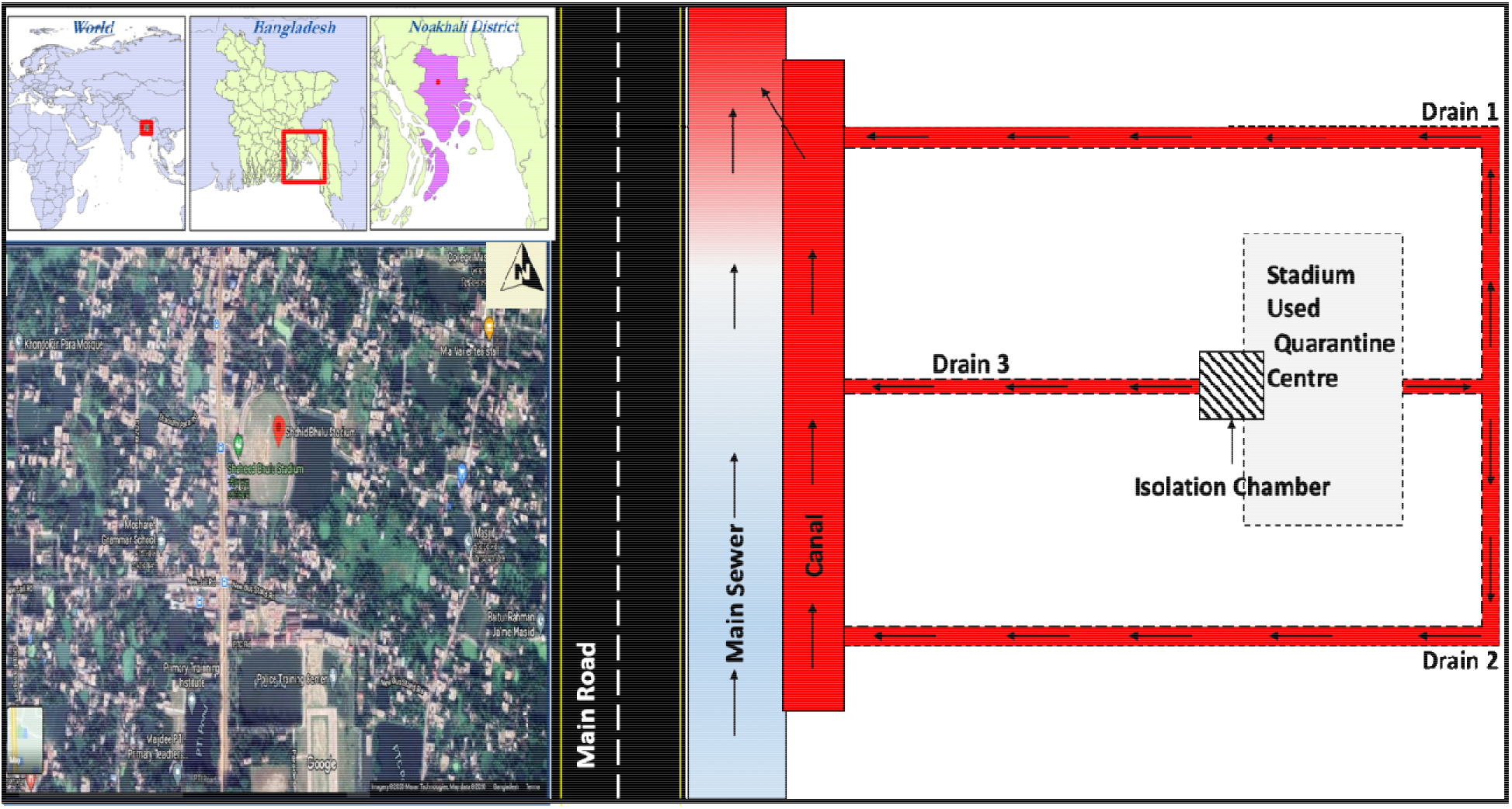
Study area location depicting the isolation camp established at the Shaheed Bhulu
Stadium at Noakhali along with the schematic diagram of sampling drains.

In order to understand the distance impact on genetic loading along the drains, we collected samples at various distances i.e. 100m, 200m, 300m, and 400m as presented in **Table 1**. Specimens were aseptically collected in a 50 ml sterile falcon tube, transported in the laboratory keeping inside the ice-box, refrigerated at 4°C during preparatory activities, and were analyzed on the same day. Sterile falcon tubes for sampling with identical blanks were analyzed to determine any possible contamination during the transport. All analyses were done at the Microbiology Laboratory of the Department of Microbiology, Noakhali Science and Technology University (NSTU), Bangladesh.

**Table 1.**
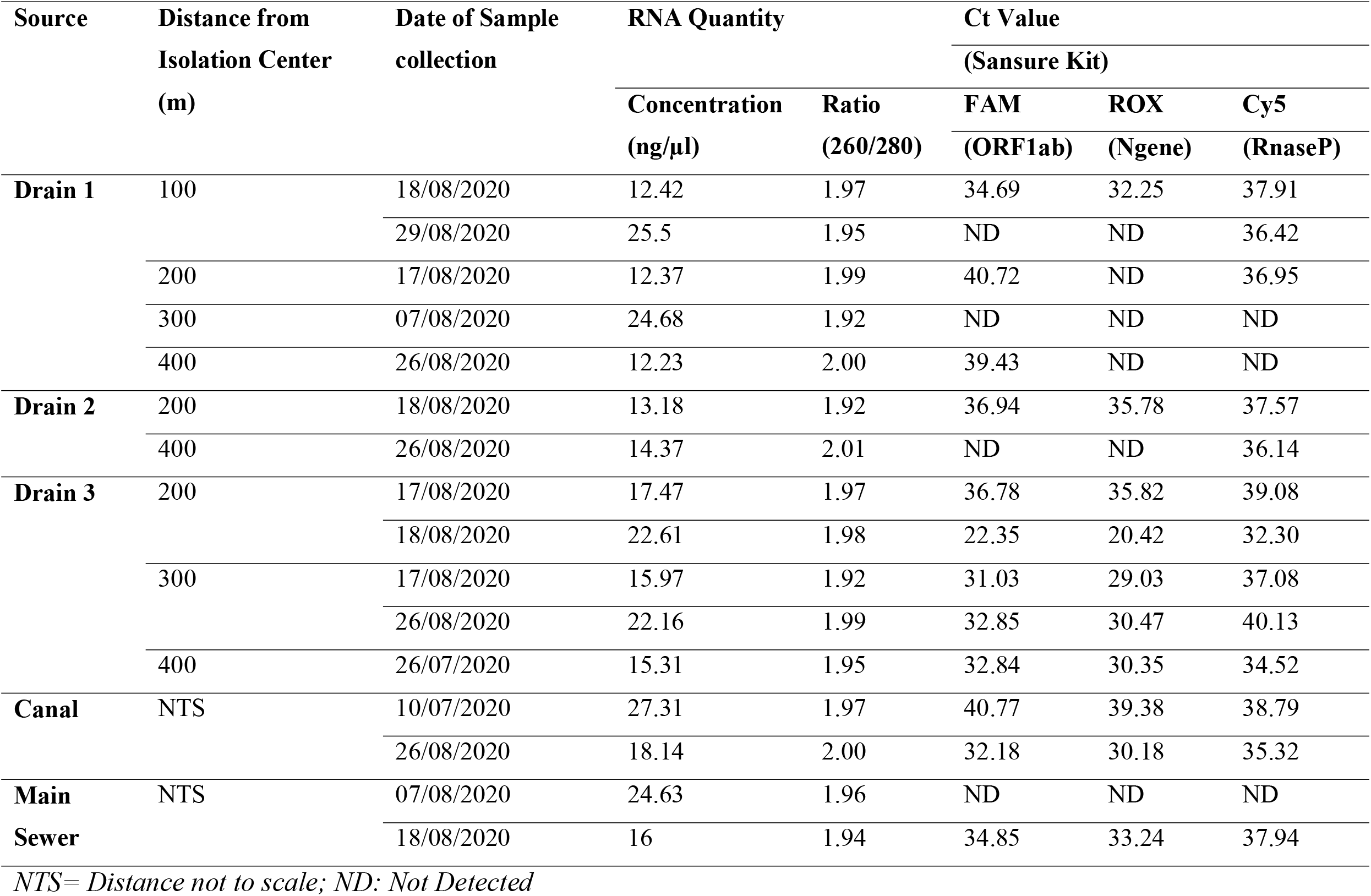
Summary of the results of amplification cycles (Ct) of various sampled wastewater along with the distance of sampling in the vicinity of isolation center with 50 days temporal resolution i.e. in between 10^th^July and 29^th^ August 2020.

### 2.2 Sample preparation, and procedure for the RNA extraction and concentration

We almost followed the same procedure of extraction described in Kumar et al. (2020). Briefly, sewage samples (50 mL) were centrifuged (Thermo Scientific) at 4500×g for 30 min followed by filtration of supernatant using 0.22-micron filters (Himedia). Further, each sewage filtrate was concentrated using the polyethylene glycol (PEG) method. In this method, PEG 6000 (80 g/L) and NaCl (17.5 g/L) were mixed in 25 ml filtrate, which was then incubated at 17°C in 100 rpm shaking for overnight. The next day, the mixture was centrifuged at 13000×g for 90 min. After centrifugation, the supernatant was discarded and the pellet was resuspended in 300 μL RNase free water. This was further used as a sample for RNA isolation. RNA isolation was carried out using a commercially available kit according to the manufacturer’s instructions. RNA concentrations were measured by NanoDrop (Thermo Scientific™ NanoDrop 2000 and 2000c, BioRad) and were stored at –70 °C until further use.

### 2.3 RT-PCR Analysis

RNAs were analyzed for the detection of SARS-CoV-2 by RT-PCR (CFX96, BioRad) using the Sansure RT-PCR kit (Sansure Biotech Inc., China). Technical procedures carried out as described in the product manual and interpretations of results were analyzed as instructed in the manual. In brief, we set the samples layout with RT-PCR protocol covering 45 cycles containing FAM fluorescence select for ORF1ab, ROX for N gene as well as CY5 for Internal control. As quality control measures, one positive control and one negative control were also run for validation of the test procedure.

## 3. Results and discussion

We collected 16 specimens between 10^th^ July and 29^th^ August 2020 from the drain, sewage, and toilets near Shaheed Bhulu Stadium Detention Centre (DC) for COVID-19 patient at Noakhali, Bangladesh. A summary of the results of amplification cycles (Ct) of various sampled water along with the distance of sampling in the vicinity of isolation center with 50 days temporal resolution i.e. in between 10^th^ July and 29^th^ August 2020 has been presented in **Table 1** and amplification plots obtained through RT-PCR illustrating temporal variation through Ct value in various sampling drains, (a) Drain 1 (b) Drain 2 (c) Drain 3, and (d) Main sewer is presented in **Figure 2**.

**Figure 2.**
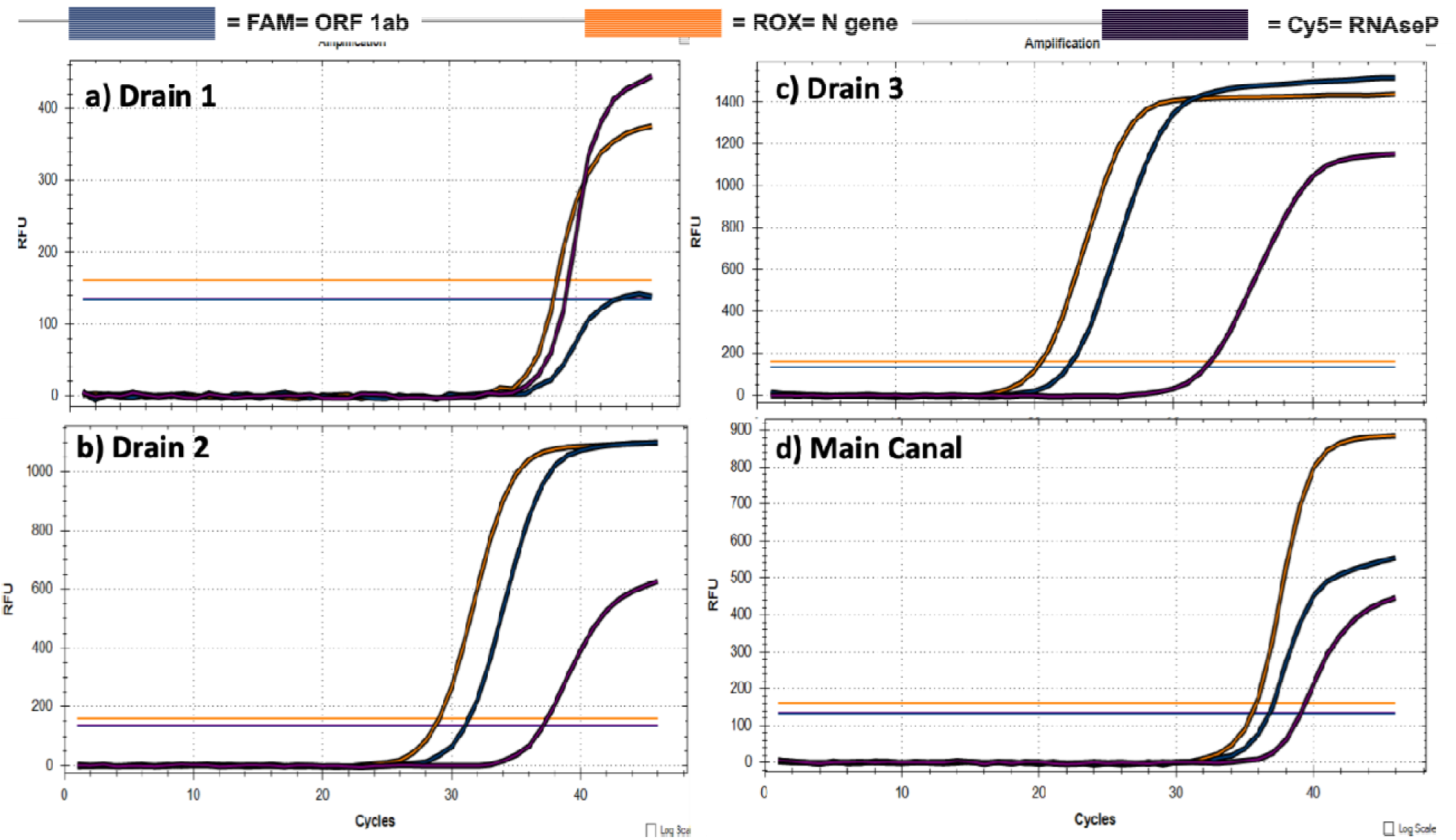
Amplification plots obtained through RT-PCR illustrating temporal variation through C_t_ value in various sampling drains.

**Table 1** summarizes the Ct values obtained during the monitoring which ranged between 20.42 to 39.38 for N genes, and 22.35 to 40.72 for ORF1ab genes, implying a huge variation in gene copies of SARS-CoV-2. Interestingly as their lowest and highest values belong to corresponding samples of the same date i.e. 17^th^ and 26^th^ August, it seems sewer systems played a critical factor WWSoC-19. Other than this anomaly 17^th^ August samples exhibited higher loading of genetic material than 29^th^ August 2020, while the number of patients in the containment remained the same during the monitoring period. This emphasizes on the fact that just becoming COVID-19 positive is not a measure of the viruses shedding by the infected person, but perhaps the state of infection matters. It is easy to speculate that with each day passing owing to aggressive testing and capacity building of carrying out the tests, early detections of COVID-19 positive people were happening, and thus probably it is reflecting on the genetic load. Moreover, temporal environmental variations due to huge rainfall with temperature and humidity fluctuations along with inadequate sewage treatment might have significant impact on the quantitative variations of SARS-CoV-2 viral genetic material.

As far as different characteristics of sampled drains are concerned, as depicted in **Figure 2**, drain 3 seems carrying the heavier RNA load of SARS-CoV-2 followed by drain 2 and drain 1. Although dissimilarities observed between the primary drainage line i.e. drain samples and secondary drainage system i.e. canal and tertiary drainage system i.e. the main sewer, however, trend of higher genetic material loading in the secondary and tertiary system was also found on twilight tenure. This is a unique finding where gene accumulation has been observed instead of the general expectation of dilution in the larger sewer system. The probable reasoning, other than the accumulation of loading from various drains of the isolation centre, in support of this observation can be the additional contribution of RNA excreted from the asymptomatic patient as well as yet to be diagnosed people into the sewer system.

We employed the distance factor in our sampling strategies and the results are presented in **Figure 3**. While drain 3 in general showed the increase in the ORF1ab and N genes, drain 1 was not showing a consistent trend in genetic material loading of SARS-CoV-2 along with the distance. We tend to propose that probably distance in meters is not likely of a critical factor capable of producing a trend.

**Figure 3.**
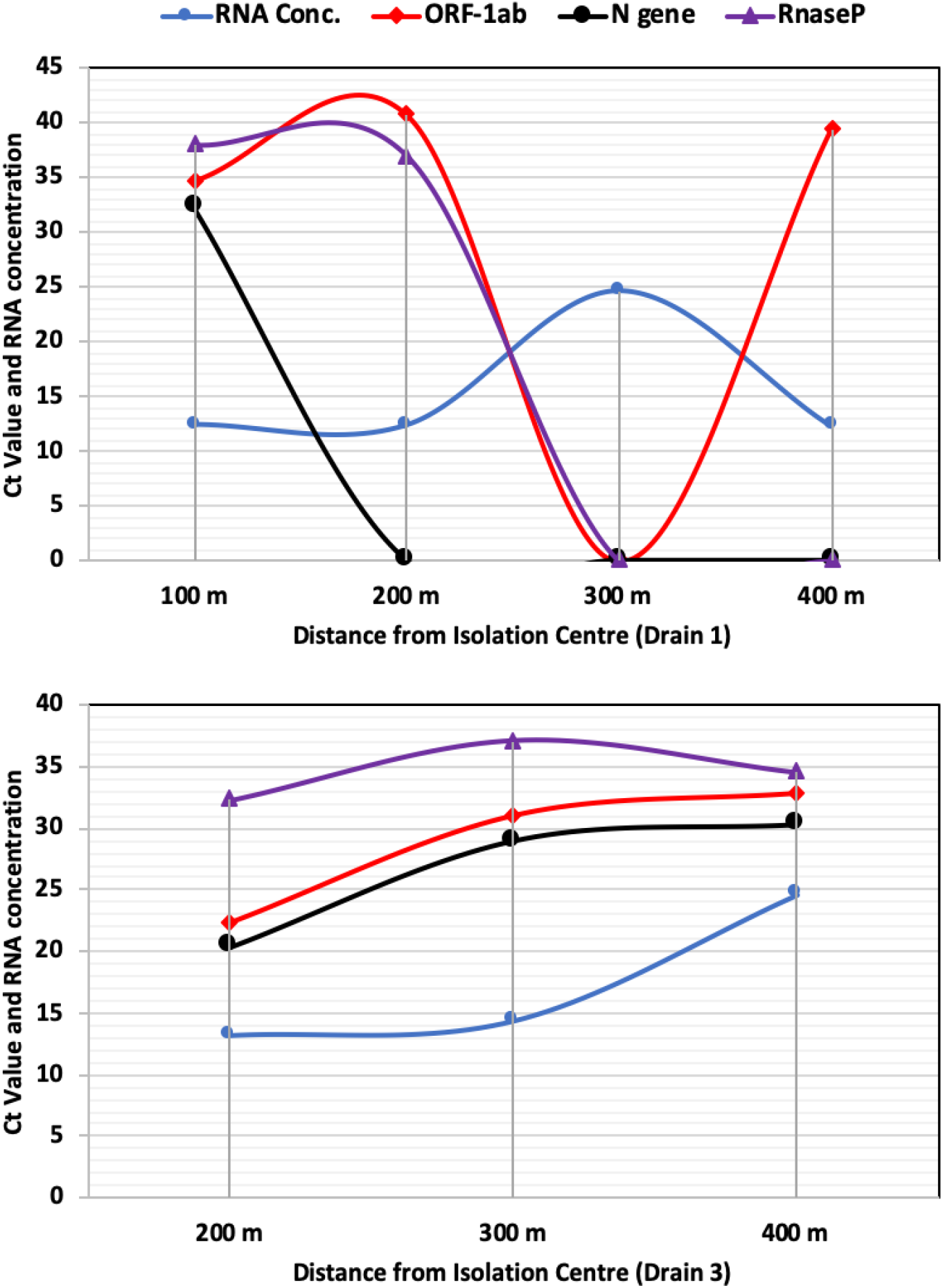
Trend in genetic material loading of SARS-CoV-2 along with the distance.

**Table 2** shows a comparative analysis of monitoring period, percentage of positive samples detected, and range of Ct-value along with their reference (Ahmed et al., 2020; Balboa et al., 2020; Kocamemi et al., 2020; Kumar et al., 2020; Medema et al., 2020; Or et al., 2020; Rimoldi et al., 2020; Wu et al., 2020b; Wurtzer et al., 2020). One finding stands out on this comparison is the Ct value of 20.42 which corresponds to much higher genetic material loading of SARS-CoV-2 than any other study reported. This may be because we sampled in the vicinity of the isolation centre, while other studies compared in **Table 2** have reported the values from the wastewater treatment plant.

**Table 2.** Comparative positive sample and the range of Ct value reported of ORF-lab genes of SARS-CoV-2 in the wastewater of various countries.

| Country | Period of Examination | Positive sample from total samples | Ct | Reference |
| --- | --- | --- | --- | --- |
| Italy | 14/04/2020 - 22/04/2020 | 4(12) | - | (Rimoldi et al., 2020) |
| Spain | 06/04/2020 - 21/04/2020 | 7(7) | 33.61 – 39.60 | (Balboa et al., 2020) |
| Turkey | 21/04/2020 - 25/04/2020 | 7(9) | 34.67 – 39.54 | (Kocamemi et al., 2020) |
| Netherlands | 05/02/2020 - 16/03/2020 | 10(24) | - | (Medema et al., 2020) |
| France | 05/03/2020 - 23/04/2020 | 100 % of samples | - | (Wurtzer et al., 2020) |
| Australia | 20/03/2020 - 01/04/2020 | 2(9) | 37.5 - 39 | (Ahmed et al., 2020) |
| U.S.A. | 08/01/2020 - 25/03/2020 | 10(12) | 33.87 - 38.39 | (Wu et al., 2020a) |
| India | 04/05/2020 - 12/06/2020 | 100 % of samples | 32.65 – 34.18 | (Kumar et al., 2020) |
| Israel | 10/03/2020 - 21/04/2020 | 10(26) | 32.76 – 38.5 | (Or et al., 2020) |
| **Bangladesh** | **10/07/2020 - 29/08/2020** | **12 (16)** | **20.42 – 40.77** | **(Present study, 2020)** |

Overall, we successfully detected ORF1ab and N protein genes from the wastewater samples of Bangladesh, which is for the first time, reported the data from the containment centre. Our study provides an opportunity to produce a realistic coefficient in the future for the conversion of genetic material loading with the number of infected people in the community.

Further, referring to the limitations of the present study, surrogate samples were sent to other laboratories to check how precisely the Sansure kit data matches with others. We also tested the filtration methods to check with the efficiency of the PEG method used in this study. Although, Hata and Honda (2020) reported a high efficiency of the PEG method in Japanese wastewater, and the same has been found true by Kumar et al. (2020) while testing the sample concentration efficiency between PEG and filtration. While lack of supply always poses critical challenges in any research, during lockdown we could not find a supply of MS2, as used by Kumar et al. (2020) or could perform the whole process control (WPC) together with MPC as recommended by Haramoto et al. (2020). We had to use a swab sample of a symptomatic person as our control as indicated by CY-5 as quality control of our analyses, which makes sense and provides a low-cost control for such analyses, yet it may be controversial, less precise and at times comes negative as the case in a few samples in our case (drain 1 samples at 300 and 400m and main sewer sample on 07^th^ and 26^th^ August). By applying fluorescence quantitative RT-PCR technology, this test utilized the novel Covid-19 ORF1ab and the specific conserved sequence of coding nucleocapsid protein N gene as the target regions, which were designed for conserved sequences of the double target genes to achieve detection of wastewater samples RNA through fluorescent signal changes. The PCR detection system used the positive internal control, which monitors the presence of PCR inhibitors in test specimens by detecting whether the internal control signal is normal, to avoid a false negative result when used for human RT-PCR experiments. In our experiments with wastewater, few specimens were negative for CY5 indicated that the human gene RNase P gene was missing in the samples. Hence it has been noted that the human RNase P gene is more vulnerable to degradation than Covid-19 viral genes.

We also refrained to put here any semi-quantitative calculation of gene copies owing to lack of enough replicates, kit designed for the human sample as well as uncertainties involving RT-PCR (Stuart et al 2014) and will induct such calculation in our future study when there will be a method at hand for precise copy calculations using suitable methods. Nevertheless, the bottom line is that the patterns of obtained Ct values suggest successful detection of SARS-CoV-2 RNA from the wastewater samples in Bangladesh, and their increasing abundance in tertiary drain demonstrated that it is not difficult to employ the COVID-19 surveillance through wastewater in the sewer systems to know the community health as we probably do not need dense sampling but the major drains may be enough to use the capability of wastewater-based epidemiology in south-Asian settings.

## 4. Conclusions

While the wastewater surveillance of COVID-19 has been focused on wastewater treatment plants around the world, we have opted for drain waters monitoring in the vicinity of the isolation center, which is first of its kind. Apart from this being the first detection report of SARS-CoV-2 RNA in the wastewaters of Bangladesh, the uniqueness of the study has been the tracing the genetic load in the vicinity of the isolation center that contains almost the constant number with 200 COVID-19 patients, which takes the variable of the number of infected persons out of the equation. This has been the key feature of this study as most of the study reported worldwide has either reported total infected person in the city or country. There has been a complete lack of infected person information that is contributing to the total genetic load to the sampled wastewater. We have found about 75% of our sampled water positive based on ORF1ab gene absence or presence. However, the critical observation has been the temporal variation where small drains showed an easing of the loading of genetic load, the bigger canal, and main sewer city exhibited temporal accumulation of SARS-CoV-2 RNA. On the other hand, the distance of sampling location in meters appears to be insignificant from the perspective of wastewater surveillance of COVID-19. We intend to analyze further samples taken in June, July, and August and preserved to shed further light on the temporal variation and decay/accumulation processes of the genetic load of the SARS-COV-2.

## Data Availability

All the data used in the manuscript are primary data and has been generated through exprimental process of the present study.

## Ethics Statement

The work did not involve any human subject and animal experiments.

## Acknowledgement

We acknowledge the sincere help extended by several COVID-19 Diagnostic Lab Volunteers to name a few are: Md. Shariful Islam, Md. Sahedul Islam, MD. Amzad Hossain, Mahmudul Islam Rakib, Golam Shamdani, Amor Chandra Nath, and Md. Julker Nyne, of Noakhali Science and Technology University, Bangladesh.

